# COVID-19 Seroprevalence in Baixada Santista Metropolitan Area – Brazil

**DOI:** 10.1101/2020.08.28.20184010

**Authors:** Karina Barros Calife Batista, Marcos Montani Caseiro, Cláudia Renata Barros, Lourdes Conceição Martins, Arthur Chioro, Evaldo Stanislau Affonso de Araújo, Marcos Calvo, Luiz Alberto Amador Pereira

**Affiliations:** Departamento de Saúde Coletiva da Faculdade de Ciências Médicas da Santa Casa de São Paulo, São Paulo, São Paulo, Brazil; Faculdade de Medicina da Universidade Lusíadas, Santos, São Paulo, Brazil; Programa de Pós-Graduação em Saúde Coletiva da Universidade Católica de Santos, São Paulo, Brazil; Universidade Federal de São Paulo, São Paulo, Brazil; Universidade São Judas Tadeu, Cubatão, São Paulo, Brazil

**Keywords:** Covid-19, Seroprevalence, Brazil, SARS-Cov-2, Inequalities

## Abstract

**Background:** COVID-19 reached Brazil in February 2020 after spreading throughout Asia and Europe. The disease first struck in Brazil’s major cities, affecting high-income groups, subsequently spreading to the socially-vulnerable outlying areas. The objective of this preliminary study was to the estimate the seroprevalence of COVID-19 in the Baixada Santista Metropolitan Area - RMBS, to inform policies for prevention and preparation of primary care, hospital services, hospital beds, intensive and critical care in a bid to reduce lethality.

**Methods:** This study is the first of a four-phase cross-sectional survey to include all cities within the Baixada Santista metropolitan area. The probabilistic population-based sample was based on a 7% prevalence with a 2% delta, a 5% level of significance and 90% power, and considered the estimated population, stratified by age, gender and city. The sample was randomized in each city and street selection was based on data drawn from the 2010 Brazilian census. The serology test was chosen from the available test kits approved by the National Health Agency (ANS) of the National Health Surveillance Agency (ANVISA), for the detection of SARS-CoV-2 antibodies. Information on socioeconomic characteristics, health service use, symptoms and adherence to social distancing measures was collected. All variables were expressed as absolute and relative frequencies and confidence intervals.

**Results:** The final sample comprised 2,342 respondents with a mean age of 37.78 (±19.98) years and equal gender distribution. Respondents predominantly lived in an abode with 4 or more rooms, 71.9% had an income of ≤3 minimum wages and no formal employment. Over half of the sample reported being reliant on the National Health Service (SUS) for health care, while 26.5% held private health insurance. Social distancing was practiced by 61.4% of respondents who answered this question. Seroprevalence measured was 1.4% for a 95% CI (0.93-1.93) based on mathematical estimates, with a ratio of 15 actual infections for every case notified, and lethality of 0.48%.

**Conclusion:** Results revealed a flattening of the epidemiological curve in the region and high underreporting of cases.

## Introduction

COVID-19 reached Brazil in February 2020 after spreading throughout Asia and Europe (Burki, 2020). The disease first struck Brazil’s major cities, affecting high-income groups and subsequently spreading to the highly socially-vulnerable outlying areas.

Amid Brazil’s current political and social climate, the pandemic has had an even greater impact than for countries in the Northern hemisphere. This is largely due to the lack of a defined policy for testing (only hospitalized patients, i.e. more severe cases, are being tested) and to the country’s continental dimensions, stark inequalities and large vulnerable population deprived of essential basic rights (Ministry of Health, 2020).

The Baixada Santista metropolitan (RMBS) area is a coastal region situated close to São Paulo city, comprising 9 cities with a population of over 1.5 million, a large port, major petrochemical hub and extreme socioeconomic inequality. Social distancing policies have been applied in the region, yet the incidence curve has increased because the restriction measures have been challenged by the Federal government (Editorial Lancet, 2020), leaving the population confused over adherence to measures stipulated by the World Health Organization. Moreover, areas with extreme urban disorganization, such as favelas, with no basic services such as clean water and sewerage system, are hardest hit because a larger number of dwellers in the same household are unable to observe the rules on social distancing.

The objective of this population-based study was to estimate the seroprevalence of SARS-Cov-2/ COVID-19 in the RMBS and inform policies on restriction measures to contain the incidence of the disease and prepare primary care, hospital services, intensive and critical care in a bid to reduce lethality.

### Casuistic and methods

The study is designed to be conducted in 4 phases with a 15-day interval between each. In each phase, a cross-sectional study will be conducted in in the metropolitan area of Santos.

The sample population was stratified by age, gender and living conditions, drawing on the 2010 Brazilian census data (IBGE, 2010).

Sample size was initially determined using the statistical package EPIINFO – version 7.2, free software from CDC-Atlanta (USA); based on a population estimated from the last IBGE census for the RMBS area of 1,831,884, estimated rate of 7%, and acceptable margin of error of 2%, allowing randomized sampling stratified by clusters, and a design effect and clustering effect of 1 (as suggested by statistical package), yielding an initial sample size of 2432 distributed samples. The test kit for detecting antibodies was chosen from those approved by the ANVISA-National Health Surveillance Agency (Resolution 777 of 18/03/20201).

The Lateral Flow (Wondfo^R^) test kit was selected as suitable, for its availability after local validation in patients with positive RT-PCR for SARS-CoV-2 and more than 14 days of symptoms. The results were read visually within 15 mins after application and no longer than 20 mins after, and the whole field team was trained on how to use and read the tests. The tests were considered valid and positive according to detection using the control strip. Tests were applied during the same time period across the whole region.

In addition, a questionnaire on socioeconomic characteristics, use of public and private health services, living arrangements, influenza symptoms and practice of social distancing and use of masks. The questionnaire was pre-tested and the researchers applying the test were trained before study commencement.

All variables were expressed as absolute and relative frequencies with respective confidence intervals (95%CI). The numeric variables were expressed as means and standard-deviations.

The study was approved by the committee of the Lusíada University, Santos, São Paulo state, under permit no. 3.992.985. All participants signed the Free and Informed Consent form.

## Results

The final sample comprised 2,342 interviews conducted in RMBS in the 1st phase, and respondents had a mean age of 37.78 (±19.98) years, range 0-93 years.

Regarding sociodemographic characteristics, respondents predominantly lived in an abode with 4 or more rooms, 71.9% had an income of ≤3 minimum wages and most had no formal employment (Table 1).

**Table 1.** Absolute and relative frequency at 95%CI for sample. Baixada Santista Metropolitan Area.

|  | Nº | % | 95%CI |  |
| --- | --- | --- | --- | --- |
| Sex |  |  |  |  |
| Female | 1253 | 53.5 | 50.7 | 56.3 |
| Male | 1089 | 46.5 | 43.5 | 49.5 |
| Cities |  |  |  |  |
| Bertioga | 83 | 3.5 | 0.5 | 7.5 |
| Cubatão | 171 | 7.3 | 3.4 | 11.2 |
| Guarujá | 389 | 16.6 | 12.9 | 20.3 |
| Itanhaém | 108 | 4.6 | 0.6 | 8.6 |
| Mongaguá | 73 | 3.1 | 0.9 | 7.1 |
| Peruíbe | 90 | 3.8 | 0.2 | 7.8 |
| Praia Grande | 378 | 16.1 | 12.4 | 19.8 |
| Santos | 559 | 23.9 | 20.4 | 27.4 |
| São Vicente | 491 | 21.0 | 17.4 | 24.6 |
| Number of rooms |  |  |  |  |
| 1 | 22 | 0.9 | 0.3 | 1.5 |
| 2 | 92 | 3.9 | 0.1 | 7.9 |
| 3 | 255 | 10.9 | 7.1 | 14.7 |
| ≥4 | 1973 | 84.2 | 82.6 | 85.8 |
| Family Income |  |  |  |  |
| ≤1 minimum wage | 674 | 28.8 | 25.4 | 32.2 |
| 2-3 minimum wages | 1009 | 43.1 | 40.0 | 46.2 |
| ≥4 minimum wages | 659 | 28.1 | 24.7 | 31.5 |
| Has anyone in your family lost part of their income |  |  |  |  |
| Yes | 667 | 28.5 | 25.1 | 31.9 |
| No | 1675 | 71.5 | 69.3 | 73.7 |
| Healthcare |  |  |  |  |
| Reliant on Public Health System | 1413 | 60.3 | 57.7 | 62.9 |
| Uses Public plus private health systems/paid private healthcare | 291 | 12.4 | 8.6 | 16.2 |
| Holds private health insurance | 621 | 26.5 | 23.0 | 30.0 |
| Uses paid private healthcare | 17 | 0.7 | 0.03 | 4.7 |
| Formal employment |  |  |  |  |
| No | 1665 | 71.1 | 68.9 | 73.3 |
| Yes | 677 | 28.9 | 25.5 | 32.3 |
| Practicing social distancing |  |  |  |  |
| No | 859 | 38.6 | 35.3 | 41.9 |
| Yes | 1368 | 61.4 | 58.8 | 64.0 |

Table 1 also shows that over half of the sample reported being reliant on the National Health Service – SUS for health care, while 26.5% held private health insurance. Lastly, social distancing was practiced by 61.4% of respondents who answered this question.

The seroprevalence observed was 1.4% at a 95%CI level (0.93-1.93) (Table 2).

**Table 2.** Seroprevalence of COVID-19 in Baixada Santista Metropolitan Area. 2020.

|  | Nº | % | 95%CI |
| --- | --- | --- | --- |
| Negative / non-reactive | 2309 | 98.6 | 98.0 – 99.0 |
| Positive / reactive | 33 | 1.4 | 0.3 – 5.0 |
| Total | 2342 | 100.0 |  |

Drawing on mathematical estimates for the total population of the Baixada Santista metropolitan area (1,831,884), according to the SEADE 2020, for each notified case, there are a further 15 positive infections not notified.

Of the 33 individuals testing positive, 54.5% were male, 84.8% resided in an abode with 4 or more rooms, 75.8% had an income of up to 3 minimum wages, 54.5% had no formal employment, 65.6% reported practicing social distancing and 69.7% reported relying on the national health service (SUS) for healthcare.

## Discussion

The seroprevalence measured, based on antibody detection, showed that the actual number of infected individuals far exceeded the number of official notifications (Thomas et al., 2020). In addition, the results of the analysis on lethality were discrepant, yielding an estimated rate of 0.48%, while official notifications indicate 7.2% lethality, close to global rates reported. These, disparities are likely due to the fact that, in Brazil, there is no national policy in place for mass testing, where responsibility for this investment lies with the cities and states, with majority of tests currently performed in hospitalized/severe cases only. Another concerning issue with regard to low testing is the undernotification, which was estimated at 15 actual infections for every positive case, potentially leading to a scenario of mass contamination. Efforts to tackle the COVID-19 pandemic in Brazil have been hampered by limited information owing to the low serological testing capacity of only 1,597 tests per 1 million population (Wordometer, 2020)^1^. Diagnostic confirmation is limited to moderate-severe cases, preventing a true epidemiological picture and seriously distorting estimated lethality of the disease.

To date, few studies estimating the prevalence of infection by SARS-CoV-2 have been conducted in countries in the Southern hemisphere and reports in low-to-middle-income countries are scarcer still.

Given the current scenario in Brazil, the pandemic has had a particularly big impact compared to most countries, especially those situated in the Northern hemisphere. Besides lack of adequate testing, Brazil is a country of continental proportions, characterized by economic and social heterogeneity and major inequalities. A large contingent of the population is socially vulnerable and deprived of basic rights (Thomas et al, 2020). For Brazil and its subnational territories (states and cities), which are at different stages of the pandemic, population-based studies estimating the seroprevalence in different regions of the countries can be of great value, helping to guide public policies in general and those of the National Health Service (SUS).

Another important issue, given the insufficient testing in the population at large, is obtaining a more accurate figure for lethality of the pandemic of SARS-Cov2 and Covid-19 in the region. Given its geographical proximity and population characteristics, it can be safely assumed that a very similar situation is likely unfolding throughout the metropolitan region of São Paulo city. Thus, the present study is the first of its kind in the Southern hemisphere, particularly for inclusion of the large number of respondents both in **phase 1** (comprising 2, 342 interviews and the same number of tests performed in the 9 cities of Baixada Santista), and the overall sample of 10,000 individuals set to be tested and interviewed by the end of the **4-phase** study.

The economic impact of the pandemic evidenced by the empirical data, with around 1/3 losing their jobs and over half seeing a drop in monthly income of over 50%, can also guide policy on support programs that help people continue practicing social distancing which are crucial, given the low proportion of the population with positive antibodies for SARS-Cov2.

The major economic impact of the pandemic on people’s lives highlights the need for effective public policy actions targeting the most vulnerable strata of the population (as seen in Portugal and Argentina), where these should provide specific benefits for those hardest hit in terms of income loss and/or social exclusion. Despite the difficulties outlined, social distancing had been practiced by over 60% of interviewees since the need for these measures was recommended by the government and health authorities (WHO). It is also notable that individuals, despite the economic hardships faced, continue to have faith in the data available, on health professionals and scientific evidence (Editorial Lancet, 2020). The use of masks when going out has been adopted by the vast majority of the population. The study results raise the issue of the importance of accurate information via the media in general, where fake news can promote erroneous approaches to tackling the epidemic.

Another key finding was the major reliance (60%) by the population on the national health service or SUS (Sistema Único de Saúde). Taking these SUS users together with those who make combined use of the public health system and private health care, underscores the pivotal importance of supporting the healthcare services and professionals involved in tackling the pandemic. These findings portray the profile of the Brazilian population, where approximately 70% of the population is dependent on the public health system (SUS) for healthcare (Stopa et al, 2017; Macinko et al., 2015). In addition, it is important to note that when respondents had to access health service, they did so through primary health via Basic Health Units (UBS). Over 80% of interviewees did not seek health services, suggesting that the population is protecting the health services, probably considering and recognizing the importance of keeping health professionals focused on tackling the Pandemic. The need to strengthen the National Health Service was apparent, along with the need for funding, training and provision of Personal Protective Equipment (PPE) for health professionals, among others. This should be provided for the whole system, prioritizing health professionals of hospitals, besides those working primary care.

The use of protocols for attending, management and follow-up of COVID-19 cases, including those self-isolating at home, with benign evolution of the condition, thereby avoiding overload of hospital services, is also key to tackling the pandemic.

These issues are of great importance for planning management actions and devising public health policies, such as distribution and access of ICU beds, mobile health units, procurement of supplies and medications, among others.

## Conclusions and Final considerations

The present study allowed the prevalence of infection by the disease in the region to be determined and found that the number of infected individuals is around 13 times higher than the number of officially notified cases. The survey data also suggests one individual in every 69 is infected by COVID-19 and that lethality is lower than the rate calculated from notifications, which constitute mainly moderate and severe cases.

It is envisaged that conducting the further three phases planned as continuation of this study can help overcome the methodological problems and limitations encountered in the production and analysis of the empirical material, and also shed light on the epidemic behavior and effectiveness of health measures adopted in combating COVID-19.

These findings reiterate the importance of adopting and maintaining preventive measures, such as universal use of face masks by the population and, in particular, the expansion of horizontal social distancing measures imposed by the regional and local government. This advice is of special relevance in Brazil, given the position taken by the incumbent President Jair Bolsonaro, who has recommended resumption of economic activities and the non-adoption of protective measures of social distancing recommended by the World Health Organization, practiced successfully in most countries affected by the COVID-19 pandemic.

## Data Availability

All data can be requested by

http://saude.sp.gov.br/coordenadoria-de-recursos-humanos/areas-da-crh/qualidade-de-vida-do-trabalhador-da-saude/programa-de-preparacao-para-a-aposentadoria/o-programa-nas-unidades/hospital-guilherme-alvaro-em-santos

## Declarations

Not aplicable

## Ethics approval

committee of the Lusiada University, Santos, São Paulo state, under permit no. 3.992.985.

